# Epigenetic g predicts cognitive aging and incident dementia in a diverse, nationally representative sample of older adults

**DOI:** 10.64898/2025.12.23.25342931

**Authors:** Jessica D. Faul, Stacey Collins, Trey Smith, Eric T. Klopack, Colter Mitchell, Eileen M. Crimmins, Mateo P. Farina

## Abstract

**Background:** Alzheimer’s disease and related dementias (ADRD) are major public health concerns. DNA methylation (DNAm)–based biomarkers such as GrimAge and PhenoAge predict aging and health risk, but were not designed to optimize prediction of cognitive decline. *Epigenetic g*—a DNAm-derived index of general cognitive ability—is a promising marker of cognitive function that has not been assessed in a racially and socioeconomically diverse population.

**Methods:** We used data from the 2016 Venous Blood Study of the Health and Retirement Study (HRS), a nationally representative cohort of U.S. adults aged ≥51 years (N = 3575 with high-quality DNAm). Epigenetic g scores were computed using CpG weights from a BayesR+ model of general cognitive ability developed in Generation Scotland. Cognitive function was measured with a modified version of the Telephone Interview for Cognitive Status (TICS) at each interview wave; 6-year incident dementia was defined using the validated Langa–Weir algorithm. Linear regression estimated associations with cognitive scores; logistic regression estimated 4-year dementia risk. Models were adjusted sequentially for demographics, education, parental education, APOE ε4 status, and blood-based neurodegeneration markers (NfL, GFAP, Aβ42/40, pTau181).

**Results:** Higher epigenetic g was associated with better baseline cognition (β=2.55, 95% CI 1.92–3.17) and cognition at the time DNAm was measured (β=2.30, 95% CI 1.62–2.99) after demographic adjustment. Associations attenuated but remained significant with education and parental education (β=1.23–1.89). Each unit increase in epigenetic g predicted 29% lower 6-year risk of dementia (fully adjusted HR=0.71). Results were robust to adjustment for APOE ε4 and neurodegeneration biomarkers.

**Conclusions:** Epigenetic g is a scalable, blood-based marker of cognitive function and dementia risk that adds predictive value beyond demographics, socioeconomic indicators, APOE, and neuropathology. Its validation in a diverse, nationally representative U.S. cohort underscores its potential for early risk profiling and for research on social determinants of cognitive aging in cross-national samples.

## BACKGROUND

Alzheimer’s disease and related dementias (ADRD) are a growing public health concern that affects millions worldwide. To identify at-risk groups and individuals, researchers have collected and analyzed biomarkers of cognitive impairment. While several studies have incorporated biomarkers from multiple biological systems (i.e., cardiovascular, metabolic, endocrine, etc.), one of the most promising areas of research has been the identification of molecular changes – namely DNA methylation (DNAm) – as predictors of age-related outcomes, including cognitive function and impairment.

The promise of the DNAm-based biomarkers has led to a proliferation of health measures predicting mortality (i.e. GrimAge), multisystem dysregulation (i.e. PhenoAge), and system specific health outcomes (i.e. inflammation, immune functioning, etc.). Many of these markers are associated with cognitive functioning and impairment in older adulthood [1, 2], providing researchers opportunities to examine individual and group-based differences in risk along with examining the association with social and environmental exposures across the life course (e.g. educational attainment) [3, 4]. However, as these markers were not developed to predict cognitive functioning, DNA methylation sites that are associated with cognitive function or decline may be excluded from these predictive algorithms. As a result, existing measures may not be optimized to predict cognitive functioning or health risks in later life. Thus, developing a DNAm-based biomarker tailored to predict cognitive functioning or decline holds substantial promise for advancing the study of cognitive aging. Such a measure could enhance our ability to not only detect individuals or groups who are at greater risk of cognitive impairment years or decades before the onset of systems but also uncover molecular pathways that underlie social disparities in brain health.

Prior work has constructed epigenetic based algorithms of cognitive functioning. One notable approach is the development of “epigenetic *g,*” an aggregate measure derived from DNA methylation sites associated with general cognitive ability (*g*) in adulthood, conceptually paralleling the psychometric construct of *g* in cognitive psychology [5]. The concept of *g* is thought to represent mental capacity, broadly, which influences performance on a wide array of cognitive tasks like problem-solving, reasoning, and learning [6]. However, it also likely reflects inputs like education and cognitive stimuli across the life course. Using the Generation Scotland Study, researchers developed an epigenetic measure of cognition based on several domains of cognitive performance that was replicated and predictive of cognitive functioning in two external samples of adults from the Lothian Birth Cohort Study. Other studies have used this measure of epigenetic *g* to evaluate social differences across the life course, but with mixed results [7, 8]. While this existing epigenetic *g* measure represents an important advancement, this measure was derived in a relatively homogeneous, midlife cohort with limited racial, ethnic, and socioeconomic diversity, raising questions about its generalizability to populations in other contexts. DNA methylation and cognitive ability are sensitive to social and environmental exposures throughout life that vary significantly across national and demographic contexts, including cumulative disadvantage, educational quality, and structural racism—factors that also shape cognitive health disparities. As such, it is unknown whether the structure and correlates of epigenetic *g* replicate in older, more diverse populations.

Using the Health and Retirement Study (HRS)—a nationally representative panel of U.S. adults over age 50—we apply an epigenetic measure of cognition in a population that reflects the social, economic, and biological diversity of cognitive aging in the United States. This approach extends prior work on longitudinal disease prediction and risk profiling for dementia by integrating molecular data and life course perspectives on cognitive health and dementia risk. Specifically, we seek to examine the relationship between epigenetic *g* and educational attainment, parents’ education, cognitive function, and dementia risk to determine whether a DNA methylation-based score of epigenetic *g* provides unique insights into cognitive decline and dementia risk beyond existing genetic and biological measures.

## METHODS

### Sample

To examine the association of epigenetic g on general cognitive functioning, we used data from the Health and Retirement Study (HRS), a nationally representative, longitudinal study of U.S. adults 51 years and older. The HRS collects extensive demographic and cognitive functioning information, alongside a rich set of biomarker and genetic data [9–11]. The HRS is sponsored by the National Institute on Aging (NIA U01AG009740) and is conducted by the University of Michigan.

In 2016, the HRS conducted the Venous Blood Study (VBS). To be eligible for the VBS, respondents had to reside in the community, complete the 2016 HRS core interview without a proxy, and consent to and complete the blood draw [12]. Blood samples were collected in respondents’ homes by trained phlebotomists and processed at the CLIA-certified Advanced Research and Diagnostic Laboratory at the University of Minnesota.

The sample with DNAm measurement and assays of blood-based neurodegenerative markers was selected from 2016 HRS respondents born in 1959 and earlier who participated in the 2016 VBS. From this group, a probability sample was selected from among respondents who were 65 and older and thus eligible for the 2016 Harmonized Cognitive Assessment Protocol (HCAP) or who are eligible for a future HCAP (a random half of HRS sample members not yet 65 years old). This subsample fully represents the entire HRS sample when sample weights are used.

DNA methylation profiling was performed using the Illumina Infinium MethylationEPIC BeadChip (850K) array using DNA extracted from the buffy coat collected from blood samples in 2016. DNA samples were randomized across analytic plates by age, cohort, sex, education, and race/ethnicity along with 39 pairs of blinded duplicates. Data preprocessing and quality control were performed using the minfi package in R. A total of 3.4% of the methylation probes were removed from the final data because their detection P-value fell below the threshold of 0.01 (n = 29,431 out of 866,091). Analysis for failed samples was done after removal of detection P-value failed probes. A total of 58 samples were removed after applying a 5% cutoff. Sex-mismatched samples and any controls (cell lines, blinded duplicates) were also not included in the analyses. The final analytic dataset includes 97.9% of the originally plated samples (n = 4,018). More information is available elsewhere [13]. For this analysis, missing beta methylation values were assigned a value of 0.

The final analytic sample includes individuals with complete DNAm data, cognitive assessments from the 2016 core interview, salivary DNA, and relevant demographic information (N=3575).

### Measures

#### Cognitive Functioning

Cognitive functioning was assessed using the modified version of the Telephone Interview for Cognitive Status (TICS). The total score ranges from 0 to 27, with higher scores indicating better cognitive functioning. It consists of immediate word recall (0-10), delayed word recall (0-10), backwards counting from 20 (0-2), and serial 7s subtraction (0-5). We used cognitive functioning at baseline for each individual, usually when participants were between 51 and 56 years of age, and cognitive functioning in 2016 when the DNA methylation data was collected, on average 13.7 years later. In order to minimize the effects of item-level non-response among self-respondents, we used the imputed cognition data released by HRS [14].

#### Dementia Incidence

We classified dementia using the Langa–Weir algorithm [15], which incorporates both self-respondent and proxy information. For self-respondents, modified 27-point TICS, with scores between 0 and 6 identifying those with probable dementia. For proxy respondents, we used the 11-point proxy scale, which includes interviewer assessments (0-2), proxy-rated memory (0-4), and instrumental activities of daily living (0-5). Proxy respondents scoring 6–11 were classified as having probable dementia. This classification criteria have been validated against clinical diagnoses in the Aging, Demographics, and Memory Study (ADAMS), a sub study of the HRS with in-person neuropsychological assessments and expert adjudication of cognitive status, has been widely used in population research, and aligns with Medicare-based prevalence estimates [15, 16].

Using the Langa-Weir classification, incident dementia was defined as dementia in 2018, 2020 or 2022 among respondents who were cognitively normal or cognitively impaired but not demented (CIND) in all survey waves up through 2016.

#### Epigenetic G

*Epigenetic G* was created using the following three steps. First, we used effect size weights for CpG sites derived from a BayesR+ epigenome-wide prediction model of general cognitive ability developed in the Generation Scotland cohort [5] . Second, for each participant we standardized DNA methylation beta values (mean = 0, SD = 1) at those CpG sites. Third, we multiplied the standardized values by the published weights and summed across all sites to yield a continuous epigenetic predictor of general cognitive ability (*g*).

#### Neurodegenerative Markers

The neurodegeneration biomarkers included in this study were measured from plasma and serum using blood was collected from consenting HRS participants as part of the 2016 Venous Blood Study. Prior pilot work established the stability and validity of measuring blood-based biomarkers of neuropathology using this collection protocol [17]. Neurofilament light chain (NfL), glial fibrillary acidic protein (GFAP), amyloid-β 42:40 ratio, and Phosphorylated Tau at threonine 181 (pTau181) were all measured on the same HD-X analyzer (Quanterix Inc.). AJJ-40, AJJ-42, NfL, and GFAP were measured in plasma using the Quanterix Neurology 4-Plex E kit. Phosphorylated tau (pTau-181) was measured in serum using the Quanterix Advantage V2.0 kit. For analysis, the AB42/20 Ratio was rescaled by multiplying by 100.

#### APOE Genotyping and Polygenic Risk Scores

APOE genotyping was performed on salivary DNA samples collected between 2006-2012. TaqMan allelic discrimination SNP assays, C_3084793 (rs429358) and C_904973 (rs7412) using a 7900HT Sequence Detection System and quantified using SDS v2.1software (Applied Biosystems, Foster City, CA) were performed. Genotypes were determined using TaqMan Genotyper software. For participants for whom direct genotyping was not available, APOE status was imputed from previously genotyped array data using the Illumina HumanOmni2.5 array. Dosage data for rs429358 and rs7412 was used to assign the “best guess genotypes” to infer the APOE isoform. The salivary DNA samples were genotyped and analyzed at the Center for Inherited Disease Research (CIDR) Genetic Resources Core Facility (GRCF) and Fragment Analysis Facility (FAF) at Johns Hopkins University (https://cidr.jhmi.edu). A complete description is available from the HRS [18].

We used ancestry-specific polygenic scores for educational attainment (EDU3_SSGAC18) and general cognitive ability (GCOG2_CHARGE18) for supplementary analyses. These scores were constructed by the Social Science Genetic Association Consortium (SSGAC) and HRS using publicly available genome-wide association study (GWAS) summary statistics for educational attainment from SSGAC [19] and from the Cohorts for Heart and Aging Research in Genomic Epidemiology [20] for general cognition. Each score was calculated as the sum of risk alleles weighted by GWAS effect sizes and then standardized (mean = 0, SD = 1) within the genotyped HRS sample. More information on these measures is available elsewhere [21–23].

#### GrimAge Age Acceleration

GrimAge was developed based on the 7 DNAm surrogates of plasma proteins and smoking pack years in a two-stage procedure [24]. GrimAge age acceleration was calculated by calculating the residual that results from regressing GrimAge on chronological age. This results in a measure of age acceleration without the effect of chronological age. For GrimAge, sex is also regressed out of the measure because both age and sex were used in the identification process.

#### Sociodemographic Characteristics

Age was based on birth year. Race/ethnicity was self-reported and categorized into Non-Hispanic White/Other, non-Hispanic Black, and Hispanic. Sex was reported as male or female.

Respondent’s parental education was based on father’s and mother’s highest level of education, which was self-reported. From this information, we created a dichotomous variable indicating whether either parent had at least a high school education.

Own education was based on the highest completed year of education (0-17 years). We created three categories: 0-11 years, 12 years, and >12 years.

#### Statistical Analysis

To examine the association of epigenetic *g* and cognitive functioning as well as the sensitivity of this association after the inclusion of other key social and biomarker predictors of cognitive health, we used a series of linear regression (cognitive functioning at baseline and in 2016).

Hazard models were used to examine the association between epigenetic *g* and dementia incidence over a 6-year follow-up period. In the first model, we evaluated the association between epigenetic *g* with each cognitive functioning outcome adjusted for age, sex and race/ethnicity. Subsequent models additionally adjust for own education and having at least one parent with 12 years or more of education (Model 2), APOE ε4 carrier status (Model 3), blood-based neurodegeneration markers (Model 4), and GrimAge age acceleration (Model 5). In supplementary models, we also adjusted for polygenic scores for educational attainment and general cognition, separately by ancestry group.

Sampling weights were used to account for the sampling design and differential non-response. This approach yields estimates generalizable to the US population over 55 years of age.

## RESULTS

### Sample Characteristics

Table 1 shows the descriptive information for the analytical sample for cognitive functioning at baseline and 2016 (n=3575). The average age of the samples was 69.6 years old, and age ranged from 56 to 93 at the time of DNAm assessment in 2016. Most of the sample was non-Hispanic White (71.4%), followed by non-Hispanic Black at 15.5%, and Hispanic at 13.1%. 15.2% of the sample had less than a high school diploma, 52.5% had a high school diploma or GED, and 32.3% had more than a high school diploma. 59.9% of the sample had at least one parent with a high school diploma. At baseline, scores on the modified TICS averaged 16.8 (SD = 4; range = 1-27) and at the time of DNAm assessment the average was 15.1 ((SD = 4.3; range = 0-27). In 2016, 6.5 % of respondents were classified as having probable dementia.

**Table 1:** Descriptive Statistics of the Cognitive Function Sample and Incident Dementia Sample - HRS 2016.

|  | Cognitive Function |  |  |  | Incident Dementia |  |  |  |
| --- | --- | --- | --- | --- | --- | --- | --- | --- |
|  | n = 3575 |  |  |  | n = 2762 |  |  |  |
|  | Mean / % | SD | Min | Max | Mean / % | SD | Min | Max |
| Age in 2016 (yrs) | 69.6 | 8.9 | 56 | 93 | 69.5 | 8.7 | 56 | 93 |
| Female | 57.6% |  |  |  | 58.5% |  |  |  |
| **Race/ethnicity** |  |  |  |  |  |  |  |  |
| Non-Hispanic White | 71.4% |  |  |  | 74.3% |  |  |  |
| Non-Hispanic Black | 15.5% |  |  |  | 13.9% |  |  |  |
| Hispanic | 13.1% |  |  |  | 11.8% |  |  |  |
| **Education** |  |  |  |  |  |  |  |  |
| Less than High School | 15.2% |  |  |  | 11.7% |  |  |  |
| High School/GED | 52.5% |  |  |  | 53.5% |  |  |  |
| More than High School | 32.3% |  |  |  | 34.8% |  |  |  |
| **Parent Education** |  |  |  |  |  |  |  |  |
| No Parents ≥12th grade | 40.1% | | | | 37.5% | | | |
| 1 or more Parents ≥12th grade | 59.9% | | | | 62.5% | | | |
| **Cognitive Test Score** |  |  |  |  |  |  |  |  |
| Baseline | 16.8 | 4.0 | 1 | 27 | 17.3 | 3.6 | 7.0 | 27 |
| 2016 | 15.1 | 4.3 | 0 | 27 | 15.9 | 3.8 | 7.0 | 27 |
| **Cognitive Status 2016** |  |  |  |  |  |  |  |  |
| Dementia | 6.5% |  |  |  | 0% |  |  |  |
| No Dementia | 93.5% |  |  |  | 100% |  |  |  |
| **APOE ε4** |  |  |  |  |  |  |  |  |
| 0 |  |  |  |  | 75.6% |  |  |  |
| 1 |  |  |  |  | 21.7% |  |  |  |
| 2 |  |  |  |  | 2.6% |  |  |  |
| **Neurodegeneration** |  |  |  |  |  |  |  |  |
| NfL |  |  |  |  | 23.65 | 23.45 | 2.90 | 456.34 |
| GFAP |  |  |  |  | 104.83 | 70.70 | 5.70 | 789.10 |
| AB42/20 Ratio* |  |  |  |  | 6.57 | 2.31 | 1.10 | 58.44 |
| pTau-181 |  |  |  |  | 2.05 | 2.13 | .03 | 43.00 |
| **Incident Dementia in 6 years** |  |  |  |  |  |  |  |  |
| No Onset (n=2541) |  |  |  |  | 92% |  |  |  |
| Dementia Onset (n=221) |  |  |  |  | 8% |  |  |  |
| **Years between baseline enrollment and 2016** |  |  |  |  |  |  |  |  |
| Years | 13.7 | 6.1 | 0 | 21 | 14 | 5.9 | 4 | 21 |

\*AB42/20 Ratio has been rescaled by multiplying by 100

We also show the descriptive information for the analytic sample used to evaluate dementia incidence (also shown in Table 1; n=2762). Most demographic characteristics were similar. For this sample, neurofilament light (NfL) levels averaged 23.59 pg/mL (SD = 23.5; range = 2.9–456.34). Glial fibrillary acidic protein (GFAP) averaged 104.52 pg/mL (SD = 70.4; range = 5.7–789.1). The rescaled Aβ42/40 ratio had a mean of 6.75 (SD = 2.31; range = 1.1–58.4). Phosphorylated tau-181 (pTau-181) averaged 2.04 pg/mL (SD = 2.12; range = 0.03–43). Over 6 years of follow-up, 8% of the sample developed dementia. APOE ε4 carriage was distributed as follows: 75.6% of participants carried no ε4 alleles, 21.7% carried one ε4 allele, and 2.6% carried two ε4 alleles.

The mean epigenetic *g* was 0.03 with a standard deviation of 28.19, indicating quite wide dispersion. We evaluated the correlations between each neurodegenerative marker and epigenetic *g* – none of the correlations exceeded .06 showing low correlation levels across the measures (shown in Supplemental Table 1).

### Association of Epigenetic G with Cognitive Functioning at Baseline, Cognitive Functioning at the time of DNAm assessment (2016), and Dementia Incidence (2016-2022)

Table 2 also shows the associations of epigenetic *g* with cognitive functioning at baseline (Panel A). With adjustments for chronological age, sex/gender, and race/ethnicity (Model 1), the association remained positive (β = 2.55; 95% CI = 1.92, 3.17). Adding educational attainment and parental education (Model 2) attenuated the coefficient but the association persisted (β = 1.89; 95% CI = 1.27, 2.52).

**Table 2 -.** Association between Epigenetic g and Cognitive Performance at Baseline and in 2016 (n=3575)

| A. Baseline Cognition |  |  |  |  |  |  |  |  |
| --- | --- | --- | --- | --- | --- | --- | --- | --- |
|  | Model 1 | LCL | UCL | p value | Model 2 | LCL | UCL | p value |
| Intercept | 21.10 | 19.89 | 22.30 | <.0001 | 20.46 | 19.27 | 21.66 | <.0001 |
| Epigenetic $g$ | 2.55 | 1.92 | 3.17 | <.0001 | 1.89 | 1.27 | 2.52 | <.0001 |
| Age (Baseline) | -0.07 | -0.10 | -0.05 | <.0001 | -0.05 | -0.07 | -0.02 | <.0001 |
| Female | 0.84 | 0.60 | 1.07 | <.0001 | 1.10 | 0.85 | 1.34 | <.0001 |
| Male (ref) | --- | --- | --- | --- | --- | --- | --- | --- |
| Hispanic | -2.58 | -3.01 | -2.15 | <.0001 | -1.59 | -1.98 | -1.20 | <.0001 |
| Non-Hispanic Black | -2.27 | -2.68 | -1.85 | <.0001 | -2.05 | -2.39 | -1.70 | <.0001 |
| Non-Hispanic White/Other (ref) | --- | --- | --- | --- | --- | --- | --- | --- |
| High School/GED | . | . | . | . | -1.24 | -1.51 | -0.98 | <.0001 |
| Less than high school | . | . | . | . | -3.09 | -3.50 | -2.67 | <.0001 |
| More than high school (ref) | . | . | . | . | --- | --- | --- | --- |
| No Parent Education ≥ 12 | . | . | . | . | -0.36 | -0.64 | -0.09 | 0.008 |
| Parent Education ≥ 12 (ref) | . | . | . | . | --- | --- | --- | --- |

B. 2016 Cognition
|  | Model 1 | LCL | UCL | p value | Model 2 | LCL | UCL | p value |
| --- | --- | --- | --- | --- | --- | --- | --- | --- |
| Intercept | 26.93 | 25.88 | 27.98 | <.0001 | 25.50 | 24.44 | 26.55 | <.0001 |
| Epigenetic g | 2.30 | 1.62 | 2.99 | <.0001 | 1.23 | 0.57 | 1.89 | 0.0003 |
| Age (2016) | -0.17 | -0.18 | -0.15 | <.0001 | -0.13 | -0.14 | -0.11 | <.0001 |
| Female | 0.88 | 0.62 | 1.13 | <.0001 | 0.98 | 0.73 | 1.24 | <.0001 |
| Male (ref) | --- | --- | --- | --- | --- | --- | --- | --- |
| Hispanic | -2.33 | -2.80 | -1.86 | <.0001 | -1.06 | -1.48 | -0.64 | <.0001 |
| Non-Hispanic Black | -2.36 | -2.81 | -1.91 | <.0001 | -1.82 | -2.19 | -1.46 | <.0001 |
| Non-Hispanic White/Other (ref) | --- | --- | --- | --- | --- | --- | --- | --- |
| High School/GED | . | . | . | . | -1.68 | -1.96 | -1.40 | <.0001 |
| Less than high school | . | . | . | . | -3.95 | -4.38 | -3.52 | <.0001 |
| More than high school (ref) | . | . | . | . | --- | --- | --- | --- |
| No Parent Education ≥ 12 | . | . | . | . | -0.70 | -0.99 | -0.42 | <.0001 |
| Parent Education ≥ 12 (ref) | . | . | . | . | --- | --- | --- | --- |

Table 2 also shows the results for cognitive functioning in 2016 (Panel B). In Model 1, after adjusting for chronological age, sex/gender, and race/ethnicity, epigenetic *g* is positively associated with cognitive functioning (β = 2.30; 95% CI = 1.62, 2.99). In Model 2, after adjusting for own education and parent’s education that were associated with cognitive function, the association of epigenetic G and cognitive functioning in 2016 remained statistically significant (β = 1.23; 95% CI = 0.57, 1.89).

Table 3 shows logistic regression models predicting 6-year incident dementia. After adjustment for age, sex/gender, and race/ethnicity (Model 1), each unit increase in epigenetic g was associated with approximately a 50% decrease in the risk of dementia (HR = 0.499; 95% CI = 0.496, 0.502). Further adjustment for educational attainment and parental education (Model 2) modestly attenuated the epigenetic g coefficient but it remained statistically significant associated with a 38.9% reduction in risk per unit (HR = 0.611; 95% CI = .607, .614). In Model 3, epigenetic *g* remained significantly associated with dementia risk with the inclusion of APOE ε4 status (HR = 0.679; 95 % CI = 0.675, 0.683). After including neurodegenerative markers (Model 4), epigenetic *g* continued to show a significant with reduced risk of dementia incidence (HR = 0.689; CI= 0.695, 0.693). In the final model, we adjust for GrimAge age acceleration and find that epigenetic g continued to be associated with dementia incidence risk (HR = 0.711, CI = 0.707, 0.716).

**Table 3 -.** Hazard Model Predicting 6-Year Incident Dementia (n=2762)

|  | Model 1 |  |  |  | Model 2 |  |  |  | Model 3 |  |  |  |
| --- | --- | --- | --- | --- | --- | --- | --- | --- | --- | --- | --- | --- |
|  | Hazard Ratio Estimate | Lower 95% Confidence Limit for Hazard Ratio | Upper 95% Confidence Limit for Hazard Ratio | p value | Hazard Ratio Estimate | Lower 95% Confidence Limit for Hazard Ratio | Upper 95% Confidence Limit for Hazard Ratio | p value | Hazard Ratio Estimate | Lower 95% Confidence Limit for Hazard Ratio | Upper 95% Confidence Limit for Hazard Ratio | p value |
| Epigenetic g | 0.499 | 0.496 | 0.502 | <.0001 | 0.611 | 0.607 | 0.614 | <.0001 | 0.679 | 0.675 | 0.683 | <.0001 |
| Age in 2016 | 1.141 | 1.141 | 1.142 | <.0001 | 1.136 | 1.136 | 1.136 | <.0001 | 1.137 | 1.137 | 1.137 | <.0001 |
| Female | 0.756 | 0.754 | 0.758 | <.0001 | 0.737 | 0.736 | 0.739 | <.0001 | 0.745 | 0.743 | 0.746 | <.0001 |
| Male (ref) | --- | --- | --- | --- | --- | --- | --- | --- | --- | --- | --- | --- |
| Hispanic | 2.154 | 2.146 | 2.162 | <.0001 | 1.644 | 1.638 | 1.651 | <.0001 | 1.728 | 1.721 | 1.735 | <.0001 |
| Non-Hispanic Black | 1.410 | 1.404 | 1.416 | <.0001 | 1.310 | 1.304 | 1.315 | <.0001 | 1.175 | 1.171 | 1.180 | <.0001 |
| Non-Hispanic White/Other (ref) | --- | --- | --- | --- | --- | --- | --- | --- | --- | --- | --- | --- |
| High School/GED | . | . | . |  | 1.418 | 1.414 | 1.422 | <.0001 | 1.468 | 1.465 | 1.472 | <.0001 |
| Less than high school | . | . | . |  | 2.159 | 2.150 | 2.167 | <.0001 | 2.240 | 2.231 | 2.249 | <.0001 |
| More than high school (ref) |  |  |  |  | --- | --- | --- | --- | --- | --- | --- | --- |
| No Parent Education ≥ 12 | . | . | . | | 1.046 | 1.043 | 1.048 | <.0001 | 1.054 | 1.051 | 1.056 | <.0001 |
| Parent Education ≥ 12 (ref) | | | | | --- | --- | --- | --- | --- | --- | --- | --- |
| APOE e4 0 vs 1 | . | . | . |  | . | . | . |  | 1.742 | 1.738 | 1.746 | <.0001 |
| APOE e4 0 vs 2 | . | . | . |  | . | . | . |  | 1.896 | 1.883 | 1.908 | <.0001 |
| APOE e4 0 (ref) |  |  |  |  |  |  |  |  |  |  |  |  |
| NfL | . | . | . |  | . | . | . |  | . | . | . |  |
| GFAP | . | . | . |  | . | . | . |  | . | . | . |  |
| AB42/40 ratio* | . | . | . |  | . | . | . |  | . | . | . |  |
| pTau181 | . | . | . |  | . | . | . |  | . | . | . |  |
| DNAme GrimAge Acceleration |  |  |  |  |  |  |  |  |  |  |  |  |
|  | Model 4 |  |  |  | Model 5 |  |  |  |  |  |  |  |

|  | Hazard<br>Ratio<br>Estimate | Lower 95%<br>Confidence<br>Limit for<br>Hazard<br>Ratio | Upper 95%<br>Confidence<br>Limit for<br>Hazard<br>Ratio | p value | Hazard<br>Ratio<br>Estimate | Lower 95%<br>Confidence<br>Limit for<br>Hazard<br>Ratio | Upper 95%<br>Confidence<br>Limit for<br>Hazard<br>Ratio | p<br>value |
| --- | --- | --- | --- | --- | --- | --- | --- | --- |
| Epigenetic g | 0.689 | 0.685 | 0.693 | <.0001 | 0.711 | 0.707 | 0.716 | <.0001 |
| Age in 2016 | 1.116 | 1.116 | 1.116 | <.0001 | 1.104 | 1.104 | 1.105 | <.0001 |
| Female | 0.706 | 0.705 | 0.708 | <.0001 | 0.741 | 0.739 | 0.742 | <.0001 |
| Male (ref) | --- | --- | --- | --- | --- | --- | --- | --- |
| Hispanic | 1.775 | 1.768 | 1.782 | <.0001 | 1.794 | 1.787 | 1.802 | <.0001 |
| Non-Hispanic Black | 1.016 | 1.011 | 1.020 | <.0001 | 1.003 | 0.999 | 1.007 | <.0001 |
| Non-Hispanic White/Other (ref) | --- | --- | --- | --- | --- | --- | --- | --- |
| High School/GED | 1.399 | 1.396 | 1.403 | <.0001 | 1.382 | 1.378 | 1.385 | <.0001 |
| Less than high school | 2.315 | 2.306 | 2.324 | <.0001 | 2.276 | 2.267 | 2.285 | <.0001 |
| More than high school (ref) | --- | --- | --- | --- | --- | --- | --- | --- |
| No Parent Education ≥ 12 | 1.070 | 1.068 | 1.073 | <.0001 | 1.062 | 1.060 | 1.065 | <.0001 |
| Parent Education ≥ 12 (ref) | --- | --- | --- | --- | --- | --- | --- | --- |
| APOE e4 0 vs 1 | 1.655 | 1.651 | 1.659 | <.0001 | 1.653 | 1.649 | 1.657 | <.0001 |
| APOE e4 0 vs 2 | 1.588 | 1.577 | 1.599 | <.0001 | 1.601 | 1.590 | 1.612 | <.0001 |
| APOE e4 0 (ref) | --- | --- | --- | --- | --- | --- | --- | --- |
| NfL | 1.002 | 1.002 | 1.002 | <.0001 | 1.002 | 1.002 | 1.002 | <.0001 |
| GFAP | 1.003 | 1.003 | 1.003 | <.0001 | 1.003 | 1.003 | 1.003 | <.0001 |
| AB42/40 ratio* | 1.019 | 1.019 | 1.020 | <.0001 | 1.020 | 1.020 | 1.020 | <.0001 |
| pTau181 | 1.044 | 1.043 | 1.044 | <.0001 | 1.043 | 1.042 | 1.043 | <.0001 |
| DNAmeGrimAge Acceleration |  |  |  |  | 1.014 | 1.014 | 1.015 | <.0001 |

\*AB42/20 Ratio has been rescaled by multiplying by 100

Furthermore, in Supplementary Table 2, we evaluate the association of epigenetic *g* with dementia incidence after adjusting for polygenic scores for education and cognition, separately for African and European ancestry groups. For both groups, even after adjusting for demographic characteristics, own education, parental education, APOE ε4 status, neurodegenerative markers, PGS scores for educational attainment and cognitive functioning, and GrimAge age acceleration, epigenetic *g* was associated with 6-year incident dementia. In the European ancestry group, higher epigenetic *g* was associated with lower risk of dementia. In the African ancestry group, before adjusting for educational attainment polygenic score, it was also associated with lower odds (not shown), but in the fully adjusted model, the coefficient direction became positive such that people with higher epigenetic *g* had a higher risk incident dementia (HR = 4.379; 95% CI=4.267, 4.494).

## DISCUSSION

In a nationally representative cohort of 3,575 older U.S. adults, we tested a DNA methylation–derived index of general cognition to assess its utility as a readily obtainable blood-based marker for predicting cognitive functioning and dementia risk. Overall, we show a robust association between epigenetic *g* and cognitive functioning at baseline, cognitive functioning at the time DNAm was assessed, and dementia incidence over a 6-year follow-up period. Most importantly, epigenetic *g* remained strongly associated with these outcomes after the inclusion of own education, parental education, APOE ε4 status, neurodegeneration markers, and GrimAge age acceleration. These findings highlight epigenetic *g* as a promising, scalable blood-based biomarker for identifying older adults at elevated risk for cognitive decline and dementia beyond traditional demographic, genetic, and neuropathological markers that are increasingly being collected in community and population-based studies.

Epigenetic *g* was first validated in the Lothian Birth Cohort, which consisted of adults born in 1921 and 1936 in Scotland. In that study, it predicted general cognitive ability independent of genetic predictors, demonstrating the utility of DNA methylation markers as potential indicators of cognitive health risk. Our study extended this work to a diverse, nationally representative sample of U.S. older adults born between 1923 and 1960. We find that epigenetic *g* predicted 3% of cognitive functioning at baseline. It was also highly predictive of dementia incidence. By demonstrating comparable prediction in a racially and socioeconomically diverse U.S. cohort, our study expands the generalizability of epigenetic *g* beyond the primarily Scottish samples in which it was first validated, broadening its potential application to more diverse contexts.

Additionally, this work advances the application of epigenetic *g* as a life course biomarker of cognitive functioning. Prior work has shown a strong association of epigenetic *g* to cognitive performance at early ages. Specifically, using the Texas Twin Study, a sample of approximately 1,000 children ages 8-19 years, researchers found that lower epigenetic *g* scores were associated with lower scores on tests of reasoning, comprehension, reading and math [8]. Our work shows that association between epigenetic *g* and cognitive performance can also be found in adults in midlife and older adulthood in the US. As such, epigenetic *g* may be an important biomarker of cognitive performance and/or cognitive health risks in older adulthood that can be used to investigate aging processes across the life course and evaluate individual and population level differences in cognitive health risk. Future research should investigate whether epigenetic *g* is indicative of cognitive development in early life that is carried into older adulthood, associated with cognitive change in later life, or some combination of the two processes that determine cognitive health risk in older adults.

In our analyses, epigenetic *g* remained strongly associated with cognitive outcomes even after adjusting for key social determinants such as educational attainment and paternal education. Future analysis should consider how epigenetic *g* could help clarify how early-life disadvantage, educational experience, and cumulative stress associated with low SES may contribute to disparities in cognitive aging in older adulthood. A recent study in children that evaluated change among commonly used DNA methylation markers (i.e. PhenoAgeAccel, GrimAgeAccel, DunedinPACE) found that epigenetic *g* had the highest amount of change [25]. Future work should evaluate change in older adulthood to better examine whether epigenetic g may capture meaningful increases in risk in later life associated with life course exposures.

Additionally, while recent advancements in geroscience research have led to the collection and analyses of neurodegeneration and aging-related biomarkers to examine cognitive health in later life, epigenetic *g* was a robust predictor of cognition and incident dementia over and above well-established neurodegenerative biomarkers such as NfL, GFAP, Aβ42/40, and pTau-181 as well as APOE ε4 genotype status, suggesting that it may be a valuable and complementary biomarker of cognitive aging. As such, it may show promise as a molecular indicator capable of distinguishing between individual- and population- level risk for cognitive impairment years before the development of cognitive impairment symptoms in addition to what is commonly collected. Additional longitudinal studies should test whether changes in epigenetic *g* precede cognitive decline and whether modifying life-course risk factors can alter this signature.

DNAm surrogates, such as epigenetic *g*, allow for the investigation of exposures even if those specific exposures were not directly measured in the cohort. In our case, the HRS does not have a direct measure of cognitive “g” in its broadest sense. However, there are many examples of DNAm surrogates predicting diseases and outcomes better than the original (measured) biomarkers as has been shown with traditional cardiometabolic risk factors and cardiovascular disease [26], smoking and lung disease [27], and a DNAm surrogate for C-reactive protein predicting brain injuries [28]. Just as DNAm clocks, themselves a type of DNAm surrogate, are designed to predict specific outcomes like chronological age, mortality, and changes in blood-based markers and physical measures, epigenetic *g* predicts cognition and cognitive phenotypes well because that is what it was designed to do. If predicting dementia incidence or Alzheimer’s Disease is an end goal, it is possible that we could refine a DNAm surrogate to optimize prediction of those outcomes. One can imagine a DNAm surrogate of neurodegenerative markers, brain imaging, and other cognitive disease phenotypes outperforming epigenetic *g* in the prediction of dementia risk.

While our study leverages a large, ethnically diverse, population-based cohort with integrated social, genetic, and blood-based neurodegeneration biomarkers and longitudinal follow-up, we acknowledge some limitations. First, our ability to determine incident dementia is based on survey-based cognitive performance measures and not a clinical assessment and diagnosis.

However, this method has been used elsewhere, was validated using an expert panel with a more extensive neuropsychological battery of tests, and produces similar results to those found in Medicare claims [15, 16, 29–31]. Second, the cognitive performance tests in the HRS include a limited set of cognitive domains. Epigenetic *g* has been shown to be related to performance across a wide spectrum of cognitive domains in prior work [8]. Future work with better measurements across multiple cognitive domains should evaluate the association in older adults.

## CONCLUSION

Altogether, this work demonstrates the utility of DNA methylation-based predictors of cognitive function for improving risk assessment of cognitive health risk in a racially and socioeconomically diverse sample of older adults. By extending the epigenetic *g* measure beyond its original validation in Scottish cohorts, we provide evidence that this marker can be applied to diverse studies outside of its initial discovery context. Importantly, the portability of epigenetic *g* across populations has implications for cross-national research where differences in cognitive tests batteries and administration can make direct comparisons challenging [32]. Projects aiming to measure and identify cognitive impairment risk in older adults across high-, middle-, and low-income countries could benefit from incorporating DNAm-based measures when conventional tests are difficult to standardize. Moreover, epigenetic-based cognitive scores may be particularly valuable during periods of rapid neurodegeneration or decline when traditional or survey-based testing becomes challenging. Lastly, unlike genetic variants, DNA methylation is modifiable and responsive to contextual influences across the life course, creating opportunities for prevention and intervention. Future research should investigate the behavioral, social, and environmental factors that shape DNAm profiles of epigenetic *g* to better identify intervention targets for promoting cognitive health.

## LIST OF ABBREVIATIONS

ADRD -: Alzheimer’s disease and related dementias
DNAm –: DNA Methylation
HRS -: Health and Retirement Study
VBS -: Venous Blood Study
TICS -: Telephone Interview for Cognitive Status
CpG -: Cytosine-phosphate-Guanine
ADAMS -: Aging, Demographics, and Memory Study
NfL -: Neurofilament light chain (NfL)
GFAP -: Glial Fibrillary Acidic Protein
Aβ42/40 -: Amyloid-β 42:40 Ratio
pTau-181-: Phosphorylated Tau at threonine 181
APOE -: Apolipoprotein E
SES -: Socioeconomic

## DECLARATIONS

### Ethics approval and consent to participate -

Ethical approval for this research was obtained by the University of Michigan Health Sciences/Behavioral Sciences Institutional Review Board (IRB-HSBS). The HRS also has IRB approval for all contact and data collection from HRS participants from the UM IRB-HSBS.

### Consent for publication -

Not Applicable

### Availability of data and materials -

The datasets generated and/or analyzed during the current study are available online from the Health and Retirement Study: https://hrs.isr.umich.edu/

### Authors’ contributions

JDF - Conceptualization, Methodology, Writing—original draft, Funding. SC - Formal analysis, Methodology, Writing—review & editing. TS - Formal analysis, Methodology, Writing—review & editing. ETK - Methodology, Writing—review & editing. CM - Methodology, Writing—review & editing, Funding. EMC - Methodology, Writing—review & editing, Funding. MPF - Conceptualization, Methodology, Writing—original draft.

### Acknowledgements

The authors thank Zainab Masood for administrative assistance and Bharat Thyagarajan and Advanced Research and Diagnostic Laboratory at the University of Minnesota for sample and DNA methylation array processing.

